# COVID-19 vaccination-infection status and immunological profile from India: a case study for prioritizing at risk population for targeted immunization

**DOI:** 10.1101/2024.02.14.24302808

**Authors:** Deepika Gujjarlapudi, Ankit Mittal, Vidyavathi Devi Gajapathi Raju, Sadhana Yelamanchili Veturi, Rupjyoti Talukdar, Rupa Banerjee, Nitin Jagtap, Sannapaneni Krishnaiah, Namburu Veeraiah, Nageshwar Reddy Duvvur

**Affiliations:** AIG HOSPITALS; Asian Healthcare Foundation

## Abstract

**Background:** The COVID-19 pandemic’s global impact was mitigated through rapid vaccine development, leading to a mix of natural and vaccination-derived immunity. Immunological profile in hybrid immunity remains less studies, especially in regions where non-mRNA vaccines were used. This study focuses on the immunological profiles and predictors of immune response in one such population.

**Methods:** This was a cross-sectional study to assess their humoral and cellular immune responses based on vaccination and infection history. Immunological assays were performed to measure antispike protein and neutralizing antibodies as well as interferon-γ release assay. Multivariable linear regression model was used to estimate predictors of immune response.

**Results:** The study revealed significant differences in immune response among participants based on their hybrid immunity status, vaccination, and infection history. Higher antibody titres and cellular responses were observed in individuals with hybrid immunity, especially those with dual pre-Omicron and Omicron infections (3326 BAU/ml, IQR: 770.25-5678.25 and 4.92 IU of IFN-γ/mL, IQR:3.74-16.98 respectively, p <0.001). Age and comorbidities such as diabetes and hypertension were associated with lower antibody levels and cellular response, while vaccination and hybrid immunity correlated with higher immune responses.

**Conclusion:** The prevalence of hybrid immunity was high, yet a substantial portion of the population lacks it, indicating the necessity for targeted immunization strategies. The findings underscore the importance of prioritizing high-risk individuals, such as elderly and individuals with comorbidities, for booster vaccinations to enhance community-level protection against COVID-19.

## Introduction

The global spread of the SARS-CoV-2 pandemic was eventually thwarted by effective vaccines, that were developed, and tested in record time. Although vaccination strategies have been effective in preventing the development of symptomatic and severe disease, a majority of the population has been infected with the virus at some point in time.(1) The has led to the emergence of hybrid immunity, a combination of natural immunity and vaccination-derived immunity, that proves to be more enduring and robust than either vaccine-derived or infection-derived immunities alone. Individuals with hybrid immunity not only have higher antibody titres but also a more robust cellular immune response against viral antigens.(2–4) Both of these factors have been independently associated with varying degrees of protection against breakthrough infections, even with newer strains. While this has been evaluated in regions where mRNA-based vaccines were predominantly used, data is lacking on BBV152 or ChAdOx1 nCoV-19 vaccines.

With time the original strain of the virus evolved into various lineages and sub-lineages, leading to the emergence of variants of interest (VOIs), variants of concern (VOCs), and variants of high consequence (VOHCs). These variants have shown the capacity to evade existing immune responses.(5,6) Therefore, it is imperative to continually study the evolution of these new strains and the immune response against them. This will help in identifying the population at risk and accordingly strategize updated vaccination programs to prevent another widespread outbreak of COVID-19 infection.

In this study, we analysed the immunological profiles and predictors of immune response in the study population based on their vaccination and infection status.

## Methods

### Study design and participants

This cross-sectional study was conducted between 21st to 31st December 2023 at AIG Hospitals, Hyderabad, India, a high volume multispecialty tertiary care centre, that was also a designated COVID-19 centre at the height of the pandemic. Consecutive participants were enrolled, and blood samples were obtained for immunological testing. Demographic details such as age, gender, and comorbidities were recorded, along with vaccination status (unvaccinated, primary vaccination series, and booster vaccination), type of vaccine received and infection status. Patients with immunosuppressed status were excluded (HIV infected not on treatment, current immunosuppressive treatment, underlying solid and haematological malignancies).

The study population was categorized into six cohorts: **Cohort I:** individuals who were unvaccinated but had a history of infection (UN), **Cohort II**: those who received the primary vaccination series (2 doses) and had no reported infection (PV), **Cohort II**I: individuals who received booster doses (3 doses) and had no reported infection (BV), **Cohort IV**: patients with a history of vaccination (1 or 2 doses) and a record of infection during the first or second waves only, **Cohort V**: patients with a history of vaccination (2 or 3 doses) and a record of infection during the omicron wave only, and **Cohort VI**: patients with a history of vaccination (2 or 3 doses) and a record of infection during the first or second waves and the omicron wave. (Figure 1). Serum samples were then analysed for humoral response in the entire population whereas cellular responses were assessed in a subgroup on the study population.

**Figure 1:**
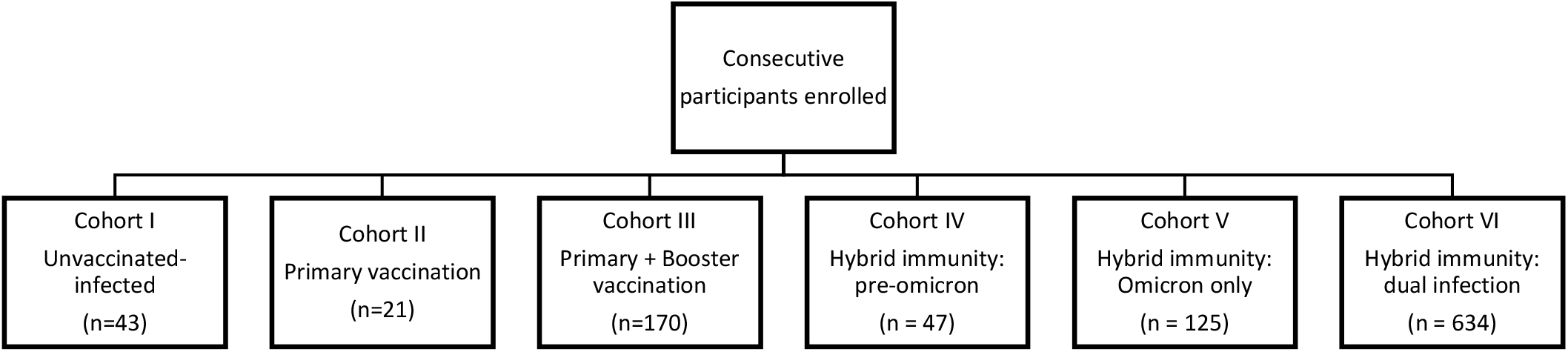
Study cohort flow diagram

The study was approved by the Institutional Ethics Committee of the AIG Hospitals (AIG/IEC-BH&R 54/12.2023-01) and written informed consent was obtained from all study participants.

### Laboratory methods

#### Assessment of humoral response

i. *Anti SARS-CoV-2 total antibodies:* The anti-SARS-CoV-2 immunoassay by ECLIA method (electro chemiluminescence immunoassay) was performed on a Cobas e601 analyser (Roche Diagnostics, Mannheim, Germany) and conducted according to the manufacturer’s instructions. This sandwich immunoassay uses a SARS-CoV-2 specific recombinant antigen representing the nucleocapsid protein, therefore is specific for SARS-CoV-2 infection. The electro chemiluminescent signal produced is compared to the cut-off signal value previously obtained with two calibrators. Results are expressed as cut-off index (COI) (negative COI < 1.0 or positive COI 1.0) for anti-SARS CoV-2 total antibodies
ii. *SARS-CoV-2 trimeric anti-spike IgG antibodies:* Serum samples were used to determine IgG anti-S1 and IgG anti-S2 antibodies by chemiluminescence immunoassay (CLIA) [DiaSorin LIASION Trimeric Spike assay (DiaSorin, Stillwater, USA)]. The IgG antibody concentration provided by the analyser is expressed as binding antibody units/ml (BAU/ml) with analytical measurement range of 4.81 to 2080 BAU/ml. The cut-off for positivity was ≥ 33.8 BAU/ml. Clinical sensitivity and specificity of this test were 98.7% and 99.5% respectively. (7)
iii. *SARS-CoV-2 surrogate virus neutralization test (sVNT):* Total immunodominant-neutralizing antibodies targeting the viral S-protein receptor binding domain were detected using a SARS-CoV-2 surrogate virus neutralization test (sVNT) assay (GenScript, Nanjing, Jiangsu, China), following the manufacturer’s instructions. The sVNT assay detects total immunodominant-neutralizing antibodies targeting the viral spike (S) protein receptor-binding domain of wild type strain Wuhan-Hu-1, Omicron B.1.1.529, or Omicron BA.2 variants of SARS-CoV-2. The percentage of serum neutralizing capacity was calculated using the equation: 1-(OD value of sample/average OD value of negative control) × 100. Values below the cut-off threshold of 30% are indicative of a negative result; The positive cut-off was a percentage of inhibition (POI) >30%, as recommended by the manufacturer.

#### Assessment of cellular immune response

*Interferon gamma release assay (IGRA):* Heparinised plasma from the study population was used to measure SARS-CoV-2 specific T-cell response by IGRA using The QuantiFERON Research Use Only platform. The QuantiFERON SARS-CoV-2 (Qiagen,Extended Pack)and Control Set were employed, pack consisted of Ag3 (Extended Pack) - CD4+ and CD8+ epitopes from S1 and S2, as in Ag2, but also immunodominant CD8+ epitopes of the whole proteome. The Control pack contains a ‘Nil tube’ which serves as the negative control and a ‘Mitogen tube’ which serves as a positive control. SARS-CoV-2 antigens that stimulate CD4+ T cells and/ or CD8+ T cells. (QFN, QuantiFERON SARS-CoV-2 Qiagen). QuantiFERON-SARS-CoV-2 was defined as positive if the IFN-γ level of S ARS CoV-2 Ag, after background subtraction, was ≥0.15 IU/mL and ≥25% of nil value as per manufacturer instructions. Following ELISA, quantitative results (IFN-γ concentration in IU/ml) were generated by subtracting the ‘Nil’ values from samples and interpolating values using an 8-parameter logistic model standard curve

#### Statistical analysis

A database was generated in MS Excel and all analyses were carried out using the Statistical Package for Social Sciences (SPSS Version 28.0 IBM, Chicago, IL, USA) software. Continuous variables were described as median value and interquartile range (IQR: 75th, 25th percentile). Continuous variables were analysed with the t test or Analysis of Variance (ANOVA) with the Bonferroni post hoc test if they were normally distributed and with Mann-Whitney or Kruskal-Wallis if they were non normally distributed. We also performed a linear regression analysis to analyse age, sex, comorbidities, vaccine type and vaccination-infection status as predictors of antibody response. A two-tailed ‘p’ value of ≤0.05 was considered statistically significant.

## Results

A total of 1040 Individuals were enrolled of which 53% were males and the median (IQR) age was 46 (26-66) years. Antibody response and sVNT was assessed in all participants, whereas, IGRA was performed in 289 participants. The baseline demographics are summarized in table 1.

**Table 1:**
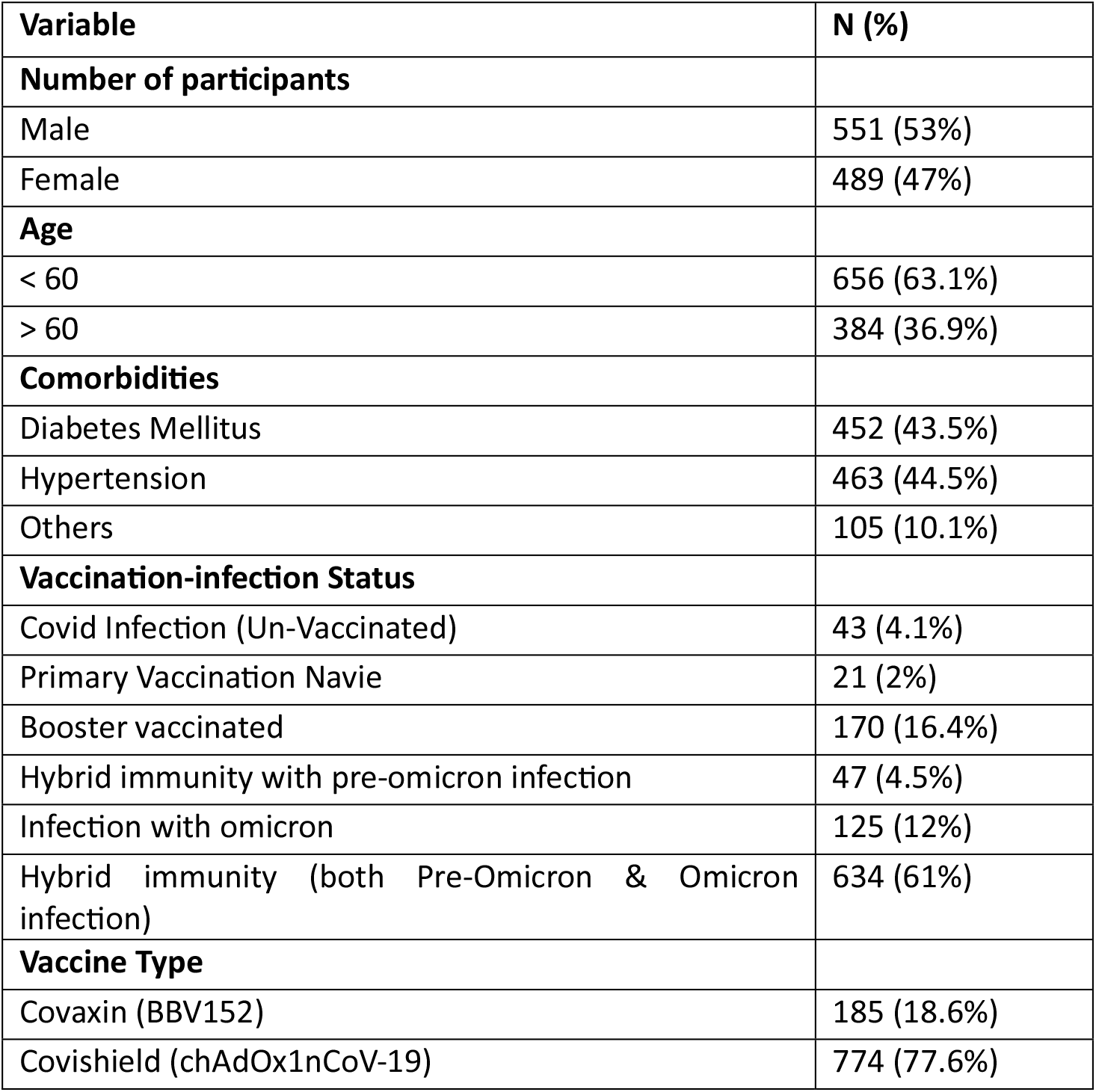
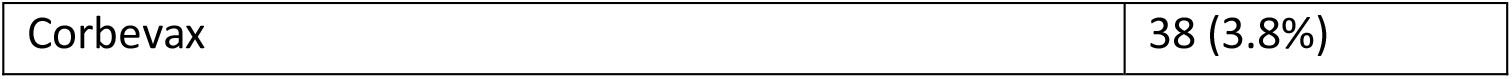
Demographic characters of the study population.

| Variable | N (%) |
| --- | --- |
| **Number of participants** |  |
| Male | 551 (53%) |
| Female | 489 (47%) |
| **Age** |  |
| < 60 | 656 (63.1%) |
| > 60 | 384 (36.9%) |
| **Comorbidities** |  |
| Diabetes Mellitus | 452 (43.5%) |
| Hypertension | 463 (44.5%) |
| Others | 105 (10.1%) |
| **Vaccination-infection Status** |  |
| Covid Infection (Un-Vaccinated) | 43 (4.1%) |
| Primary Vaccination Navie | 21 (2%) |
| Booster vaccinated | 170 (16.4%) |
| Hybrid immunity with pre-omicron infection | 47 (4.5%) |
| Infection with omicron | 125 (12%) |
| Hybrid immunity (both Pre-Omicron & Omicron infection) | 634 (61%) |
| **Vaccine Type** |  |
| Covaxin (BBV152) | 185 (18.6%) |
| Covishield (chAdOx1nCoV-19) | 774 (77.6%) |

### Humoral response profile

#### a. Seropositivity

Seropositivity rate in the unvaccinated group was 81.3% and 99.2% - 100% seropositivity was seen in individuals with hybrid immunity. The seropositivity was zero in both the primary vaccination and booster groups.

#### b. SARS-CoV-2 trimeric anti-spike IgG antibodies titres

806 (77.5%) of the participants had a hybrid immunity. The median antibody titres were significantly higher in the group with hybrid immunity compared to those with either infection or vaccination alone. Antibody titres were highest in the dual infection-hybrid immunity group (3326 BAU/ml, IQR: 770.25-5678.25) and lowest in the unvaccinated group (98.5 BAU/ml, IQR: 57.68-127.75). (Table 2) The pairwise Mann-Whitney U tests with Bonferroni correction for multiple comparisons yielded significant differences (p < 0.001) in each group (Figure 2).

**Table 2:**
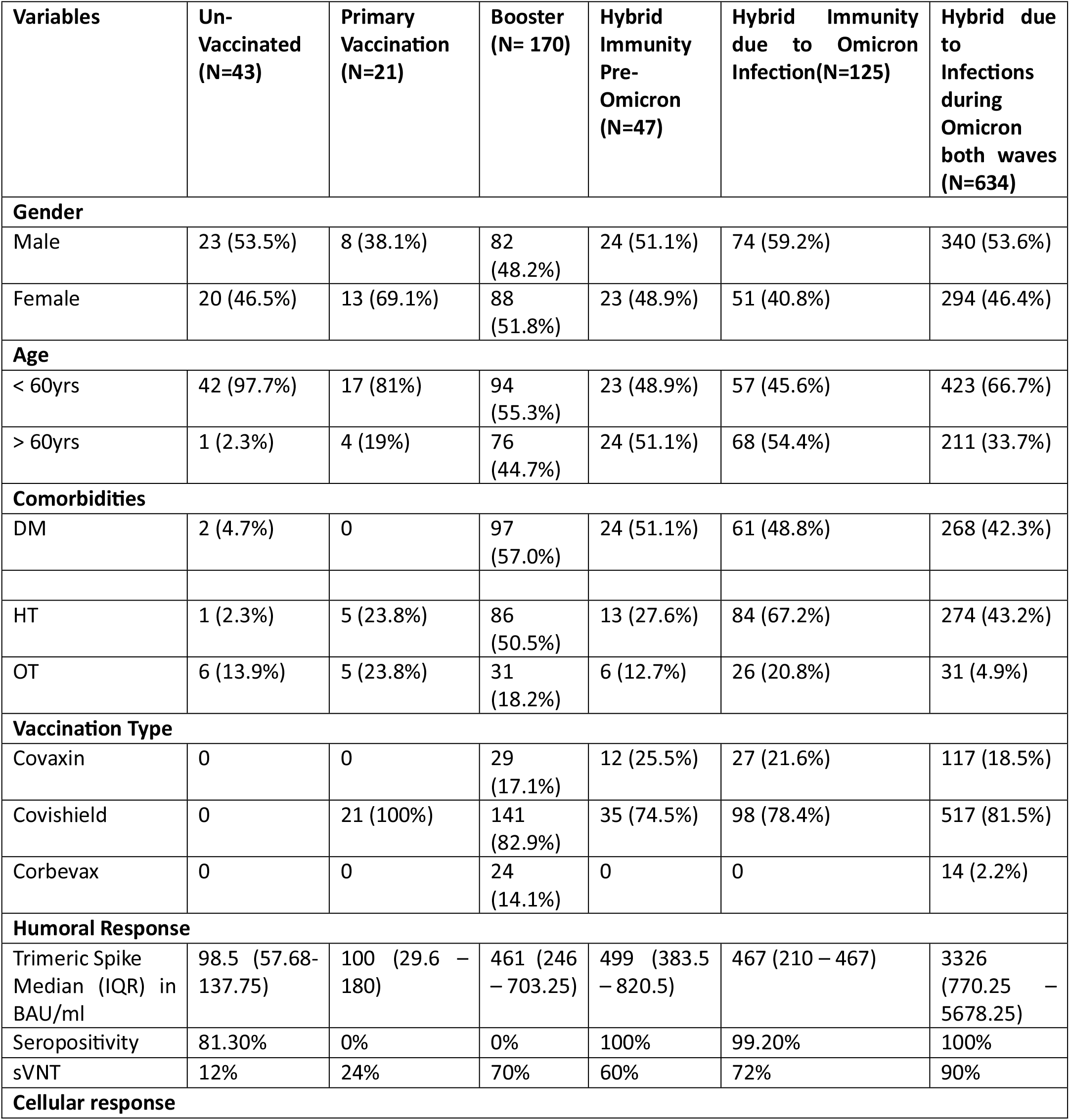
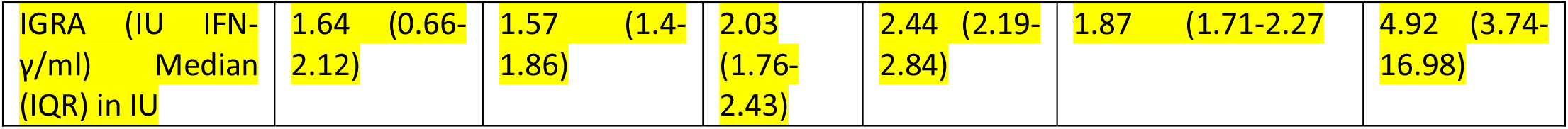

**Figure 2.**
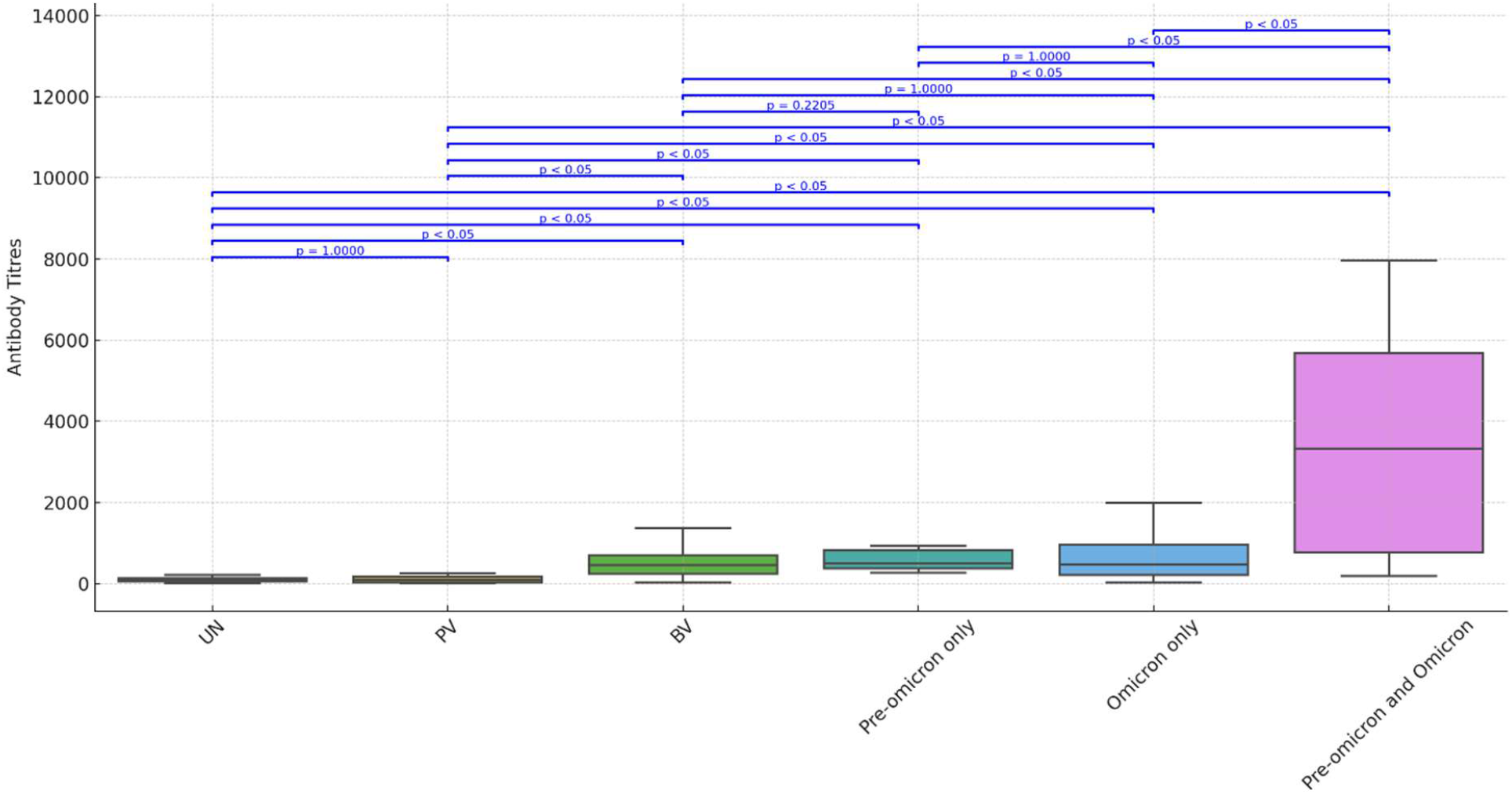
Box-whisker plot comparing the median antibody titres of the six study cohorts. The central line in each box represents the median value and the upper and lower lines represent the 3^rd^ and 1^st^ quartile respectively. The horizontal lines connect the groups with statistical comparison. A p-value of < 0.05 is considered significant. UN = unvaccinated, infected; PV = Primary vaccination without infection; BV = Booster vaccination without infection; Pre-omicron = Hybrid immunity due to preomicron infection; Omicron = hybrid immunity due to omicron infection alone; Pre-omicron and omicron = hybrid immunity due to dual infections.

Individuals younger than 60 years had a significantly higher median antibody titres (3240 BAU/ml) compared to those aged 60 and above (520 BAU/ml) (p < 0.001). Absence of diabetes or hypertension was associated with higher median antibody levels (2997.5 BAU/ml and 2985.5 BAU/ml, respectively) compared to those with these conditions (618 BAU/ml and 610 BAU/ml, respectively; p < 0.001).

On performing the multivariate analysis, individuals with hybrid immunity due to both Pre-omicron and Omicron infections had the highest antibody levels (β = 2931.44, p < 0.001). Individuals over 60 years had significantly lower antibody levels (β = -1960.19, p < 0.001). The presence of hypertension was linked to lower antibody levels (β = -739.23, p < 0.001). Unvaccinated individuals and those with only primary vaccination had lower antibody levels (β = - 1641.54 and -961.18 respectively, p < 0.001), while a booster vaccination was associated with higher levels (β = 326.83, p = 0.009).

#### c. Neutralization assay

The sVNT values against Omicron BA.2 were 12% in unvaccinated individuals, 24% in those with primary vaccination, and 70% in individuals with a booster. In cases of hybrid immunity, the values are 60% for pre-Omicron, 72% for Omicron infection, and 90% for hybrid immunity due to infections during both Omicron waves. (Figure 3). Percentage neutralization was significantly lower in the unvaccinated and primary vaccination cohorts compared to others (p< 0.001)

**Figure 3.**
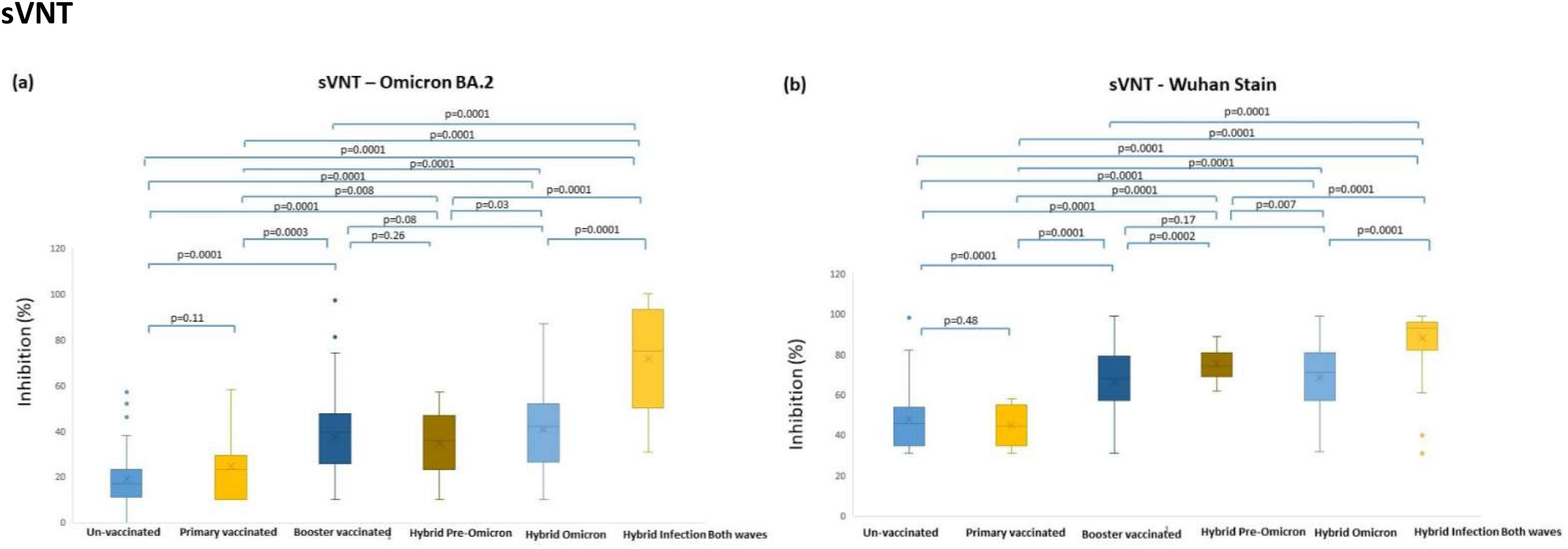
Box-whisker plot comparing the inhibition percentage on sVNT for Omicron BA.2 (a) and Wuhan strain (b) for the six different cohorts. The central line in each box represents the median value and the upper and lower lines represent the 3^rd^ and 1^st^ quartile respectively. The horizontal lines connect the groups with statistical comparison. A p-value of < 0.05 is considered significant. Hybrid Pre-omicron = Hybrid immunity due to pre-omicron infection; Hybrid Omicron = hybrid immunity due to omicron infection alone; Hybrid infection both waves = hybrid immunity due to dual infections.

### Cellular response profile

IGRA was performed for 289 participants. The cellular response was significantly different among the six groups (p<0.001). The highest median level of IGRA was observed in the hybrid immunity due to dual infection with a median value of 4.92 (IQR:3.74-16.98) IU/mL, followed by the hybrid immunity due to pre-omicron infection (2.44 IU/mL, IQR:2.19-2.84). The lowest median level was observed in the un-vaccinated group (1.64 IU/mL, IQR:0.67-2.12), followed by the primary vaccination group (1.57 IU/mL, IQR:1.40-1.86) (Table 2). (Table 2) (Figure 4). On multivariable linear regression analysis, age and presence of comorbidities were associated with poor IFN-γ response (β = -0.17; 95% CI: -0.21 to -0.14 and -2.64; 95% CI: -4.25 to -1.03 respectively, p < 0.001). Vaccination status was associated with higher IFN-γ levels (β = 3.05; 95% CI: 2.74 to 3.36, p <0.001).

**Figure 4:**
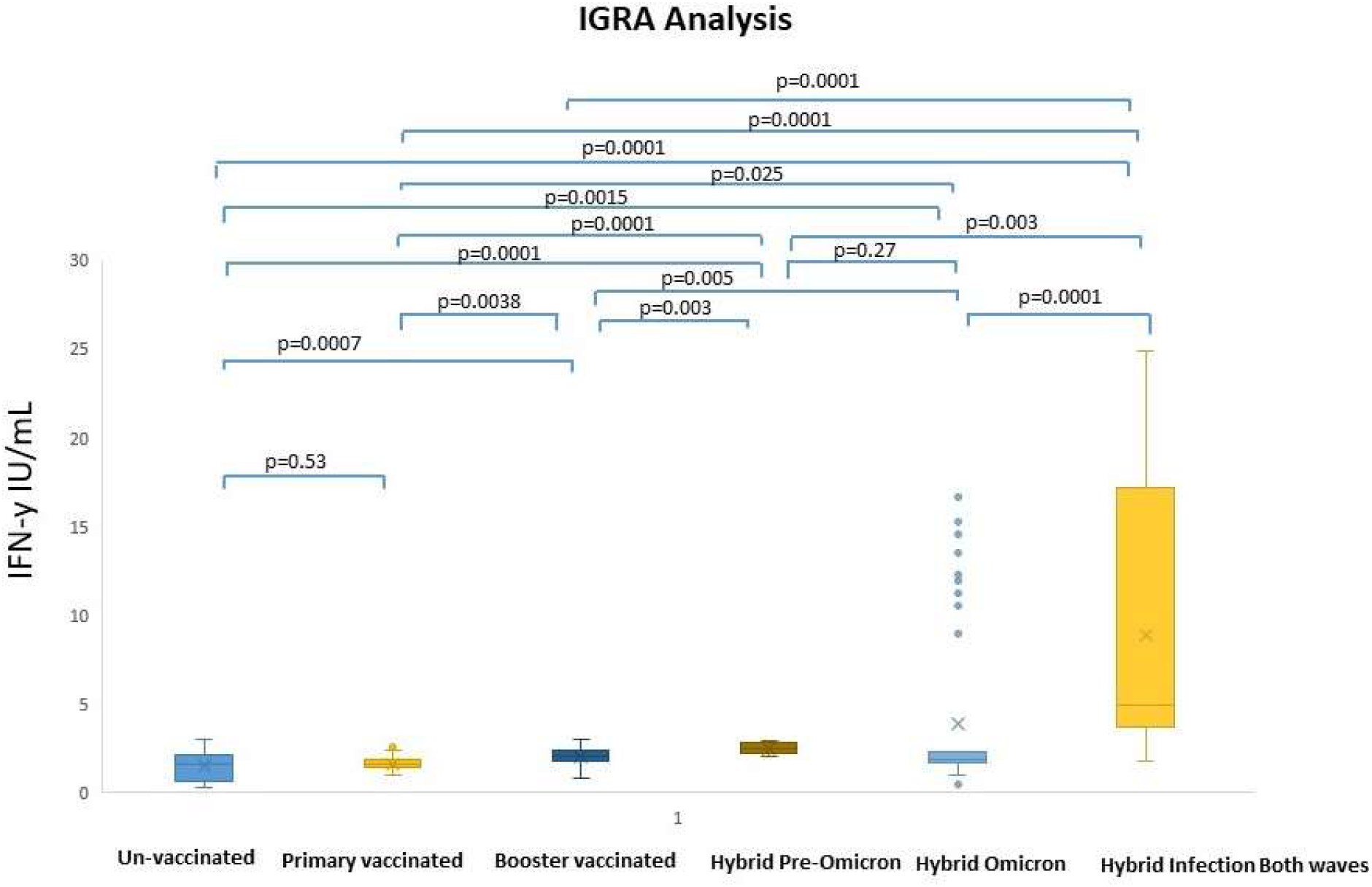
Box-whisker plot comparing the IFN-y values of the six study cohorts on IGRA. The central line in each box represents the median value and the upper and lower lines represent the 3^rd^ and 1^st^ quartile respectively. The horizontal lines connect the groups with statistical comparison. A p-value of < 0.05 is considered significant. AG3 = IU IFN-γ/ml; UN = unvaccinated, infected; PV = Primary vaccination without infection; BV = Booster vaccination without infection; Pre-omicron = Hybrid immunity due to pre-omicron infection; Omicron = hybrid immunity due to omicron infection alone; Pre-omicron and omicron = hybrid immunity due to dual infections.

## Discussion

This study assessed both humoral and cellular immune responses in a large cohort of individuals from India, where predominantly received vaccinations with BBV152 and ChAdOx1 nCoV-19 vaccines. Assessment of neutralizing antibodies and T-cell activity serve as an acceptable correlate of protection against SARS-CoV-2 infection as well as disease and therefore immunogenicity can help in strategizing immunization programmes.(8–10)

Our study demonstrates a high prevalence of hybrid immunity in the general population with a majority having more than one infection. We identified older age (specifically > 60 years) to be significantly associated with lower antibody response. This is similar to the findings by Prather et al, where increasing age was related to lower antibody titres even with mRNA vaccines. (11) Increasing age independently remain associated with lower antibody response not only in general population but also in organ transplant recipients and cancer patients.(12–14). Our findings on association between antibody response and presence of comorbidities are also consistent with existing literature.(15) Both diabetes and hypertension were associated with lower titres on univariate analysis, but only hypertension retained significance on multivariate analysis. Other comorbidities such as obesity, chronic kidney disease and immunocompromising conditions are known to be associated with poor immunogenic response and early waning of titres. (12,16)

Notably, the group with hybrid immunity due to dual infection consistently showed significant differences when compared with other groups, suggesting a strong association with higher antibody levels. Duro et al, in their study assessed the antibody response in ChAdOx-1 nCoV-19 vaccinated participants, where they found that hybrid immunity was associated with higher values as well as slower decline in the antibody titres with time.(17) Since this was a cross-sectional study, we could not assess the relationship of antibody titres with time. However, individuals in the vaccination group alone had their immunogenic stimulus much before than those with an infection. This could also partially explain the lower antibody titres in the vaccination without infection group.

Neutralization assay is generally considered a better correlate of protection as compared to total antibodies and absence of NAbs has shown the strongest correlation with mortality and delayed viral control.(18) Decay in NAbs over time is well documented and therefore high-risk groups should be boosted with additional doses of vaccines. (19) In our study, serum from individuals with no vaccination or only primary series vaccination had lower neutralization percentage compared to other groups and these groups can potentially benefit from an updated booster dose.

T cell immunity might play a crucial role in long-term protection from severe COVID-19, even in the context of emerging VOCs. Therefore, it is essential to assess the duration of cell-mediated immune protection against SARS-CoV-2.(20) In the present study, we evaluated T cell responses 20-24 months after the immunization with 2 or 3 doses of BBV152 and ChAdOx1 nCoV-19 vaccine with or without SARS-CoV-2 infection. In this study, T-cell mediated immune responses were sustained even after 24 months following the primary or booster BBV152 and ChAdOx1 nCoV-19 vaccination regimen in COVID-19-naïve individuals, with no significant differences between vaccines. We also found that the IGRA levels were significantly higher in the hybrid immunity group and booster group suggesting robust immune response. This is similar to the findings Desmecht et al, where booster vaccination and hybrid immunity was associated with a higher cellular as well as humoral response. (21) Additionally, old age was associated with a poor IFN-γ response which could be due to accelerated T-cell exhaustion. (22) Diabetes mellitus was found to be associated with poor IFN-γ response. T-cell response is known to be negatively correlated with fasting plasma glucose and glycated haemoglobin levels. (23) This correlation was however, not assessed in our study.

This is one of the few studies exploring the durability of cellular immunity after 24 months still immune response is generated to BBV152 and ChAdOx1 nCoV-19 vaccines and providing insights into efficacy against VOC and vaccination.

This study lays the groundwork for future immune-bridging investigations, allowing for a comparative analysis of the immunogenicity of new vaccine candidates, whether utilizing existing or novel platforms. This streamlined approach aims to minimize the need for extensive clinical trials, facilitating a more efficient process for timely approvals of booster vaccines.(24,25) For example, by utilizing ChAdOx1 with a 4-week spacing of doses as a reference, achieving a superiority margin of 2.6-fold in geometric mean titers (GMT) compared to ChAdOx1 vaccine, would yield a similarly high confidence level of over 80% in vaccine efficacy. (26) Gaining a better understanding of the cellular immune response to COVID-19 should be an important point of future research, in order to inform public health policies and guide targeted interventions for vulnerable populations.

### Limitations

The major limitation of this study is the exclusion of immunocompromised patients as they would also otherwise form the major ‘at-risk’ group. However, our results could still be generalized at the population level as age and two major comorbidities have been accounted for. The other limitation is the lack of microbiological confirmation Omicron infections for all the cases and there could be an under-reporting of the mild/asymptomatic infection rates. For the same reason, clinical outcomes, such as, incidence of breakthrough infection could not be analysed. Additionally, IFN-γ measurements were only available at one time-point, specifically after the third dose. This did not allow to capture the dynamic changes in immune responses over time which would have otherwise offered a more comprehensive understanding. The study is however, strengthened by a large sample size and exhaustive immunological profiling of the study population that provides valuable insights for further immunogenicity studies with new vaccine candidates.

## Conclusion

Our study demonstrates a high prevalence of hybrid immunity in the population. However, a large proportion of our population still lacks hybrid immunity. Additionally, individuals above the age of 60 years and with comorbidities are likely to have a lower antibody titre. High-risk individuals can therefore, be prioritized for immune-boosting with new vaccine candidates in a timely fashion – an undertaking that could be particularly challenging in large and populous countries like India.

Nevertheless, a continuous commitment to research is imperative to further explore and enhance our understanding of the immunological underpinnings of COVID-19 through longitudinal cohorts.

## Data Availability

All data produced in the present study are available upon reasonable request to the authors

